# Development and Validation of Interpretable Machine Learning Models for Low Birth Weight in Ethiopia: A Secondary Analysis of the Ethiopian Demographic and Health Survey

**DOI:** 10.64898/2026.08.17.26360212

**Authors:** Ashebir Mamay Gebiru, Senafekesh Biruk Gebeyehu, Serku Abate Mihret, Kumlgn Tesfa Ferede, Yimer Mamaye

## Abstract

**Background:** Low birth weight remains a primary driver of neonatal and infant mortality in Ethiopia. Machine learning models can assist early risk identification, yet clinical adoption is often limited by black box algorithms and late pregnancy predictor variables. This study aimed to develop and validate interpretable machine learning models using early pregnancy and sociodemographic features from a national survey dataset.

**Methods:** Secondary data from the nationwide Ethiopian Demographic and Health Survey were analyzed. Predictors were restricted to features accessible during early antenatal visits. Six machine learning algorithms were trained and evaluated on an independent holdout test set: Logistic Regression, Decision Tree, Support Vector Machine, Gradient Boosting, Random Forest and Extreme Gradient Boosting (XGBoost). Imbalance was addressed using synthetic oversampling on the training set. Model explainability was established through Shapley Additive exPlanations (SHAP).

**Results:** Out of 12876 births, 4249 (33%) were categorized as low birth weight / small birth size. XGBoost achieved superior predictive performance with an AUC-ROC of 0.947 (95% CI: 0.910-0.938) on the test set, outperforming lasso ML (0.8637) and standard logistic regression (0.8088). Key global predictive drivers identified by SHAP values included maternal anemia status, short inter pregnancy interval (< 18 months), low maternal BMI (< 18.5 kg/m^2), rural residence, lowest household wealth quintile and delayed or non-attendance of first trimester antenatal care.

**Conclusion:** Machine learning models trained on early pregnancy and demographic features can accurately predict low birth weight risk in Ethiopia. Integrating interpretable frameworks into primary healthcare decision support tools provides a viable strategy for early risk stratification and targeted interventions in resource-limited settings.

## Introduction

Low birth weight defined by the World Health Organization as a birth weight of less than two thousand five hundred grams regardless of gestational age (or perceived small size at birth in survey frames lacking universal clinical scales), represents one of the most critical public health challenges worldwide [1, 2]. Globally, approximately 15% to 20% of all births fall into the low birth weight category, accounting for more than twenty million newborns each year. The vast majority of these low birth weight infants are born in low and middle income countries with sub Saharan Africa bearing an exceptionally heavy burden [2]. In Ethiopia, low birth weight and small birth size remain major contributors to neonatal and infant mortality with national estimates placing the combined prevalence up to thirty three percent depending on regional disparities and socioeconomic stratification [3, 4].

Infants born with low birth weight face severe immediate and long term health consequences. In the neonatal period, these infants are at a heightened risk for severe complications including hypothermia, neonatal sepsis, hypoglycemia, respiratory distress syndrome and intraventricular hemorrhage [5]. Compared to infants of normal birth weight, low birth weight neonates are up to 25 times more likely to die during the first month of life [3]. Beyond the immediate post-natal period survivors often experience long term developmental challenges, including stunted growth, reduced cognitive performance, lower educational attainment and a heightened susceptibility to adult onset chronic conditions such as type two diabetes mellitus, hypertension and cardiovascular diseases [5].

The etiology of low birth weight in resource limited settings like Ethiopia is multi factorial, stemming from a complex interplay of maternal anthropometrics, sociodemographic factors, environmental exposures, reproductive history and maternal nutritional status during pregnancy [6, 7]. Major identified risk factors include maternal undernutrition, short stature, young maternal age, high parity; short inter pregnancy intervals, lack of formal education, rural residence and indoor air pollution from solid cooking fuels, severe maternal anemia and inadequate antenatal care utilization [4, 6]. Despite significant investments by the Ethiopian Ministry of Health to expand primary maternal and child healthcare services, timely identification of pregnant women at high risk for delivering low birth weight infants remains a substantial hurdle. In rural and remote areas where clinical diagnostic equipment such as ultrasonography is absent and healthcare workers are overburdened, traditional clinical risk assessment tools often lack both sensitivity and specificity.

Recent developments in digital health and artificial intelligence offer transformative opportunities for maternal and child health risk stratification. Machine learning algorithms can automatically identify complex nonlinear interactions among heterogeneous clinical and sociodemographic variables enabling highly accurate individual level risk predictions [3].

However, two primary barriers have historically hindered the clinical adoption of machine learning in resource constrained primary care settings. First, many machine learning studies rely on clinical variables collected late in pregnancy or during labor, limiting the window for effective preventive intervention. Second, high performing ensemble algorithms such as Extreme Gradient Boosting and Random Forest operate as opaque black box models, offering no clinical rationale for their predictions. Healthcare providers in primary healthcare units require transparent, interpretable decision support tools that explain why a particular mother is flagged as high risk, thereby enabling targeted clinical interventions [8].

To address these critical gaps, this study develops and validates interpretable machine learning models specifically designed for the early prediction of low birth weight using a nationally representative sample from the Ethiopian Demographic and Health Survey [3, 7]. By restricting candidate predictors to maternal, household and early antenatal features accessible during initial prenatal visits and by applying model agnostic explainability frameworks, this work delivers both predictive accuracy and clinical transparency to empower maternal healthcare decision making in Ethiopia [3, 8].

## Methods

### Data Source and Study Population

This study utilized secondary data from the nationwide Ethiopian Demographic and Health Survey dataset. The survey implemented a stratified, two stage cluster sampling design based on the national population census frame. In the first stage, enumeration areas were selected independently from urban and rural strata using probability proportional to size. In the second stage, a systematic sample of households was selected from each enumeration area. Data collection encompassed comprehensive face to face interviews with women aged 15 to 49 years, recording detailed information on reproductive history, maternal health service utilization, household socio demographics and health outcomes for births occurring within five years preceding the survey. The primary analytical sample for this study comprised singleton live births with recorded numerical birth weights or perceived child size at birth. Records with missing target outcome values, implausible anthropometric measures or non-singleton births were systematically excluded during data cleaning.

### Variable Selection and Feature Engineering

The primary outcome variable was low birth weight or small birth size, constructed as a binary target category where a birth weight strictly less than two thousand five hundred grams (or maternal report of very small or smaller than average size at birth) was categorized as low birth weight or small birth size and a birth weight equal to or greater than two thousand five hundred grams (or average or larger size) was categorized as normal birth weight.

Candidate predictor variables were restricted to early pregnancy features accessible to primary healthcare workers during first line antenatal contacts. Predictors were categorized into four main domains. Maternal demographic and anthropometric features included maternal age at birth, maternal educational attainment, marital status, maternal occupational status, maternal body mass index category, and maternal height. Reproductive and obstetric history features encompassed parity, birth order, inter pregnancy interval, history of preceding child mortality, desired pregnancy status and place of delivery. Socioeconomic and environmental determinants captured place of residence, geographic region, household wealth index quintile, main source of drinking water, type of toilet facility and primary cooking fuel type.

Early maternal health service features covered antenatal care attendance during the first trimester, iron folic acid supplementation during pregnancy, maternal anemia status and household media exposure.

Data preprocessing included imputing missing predictor values using K nearest neighbors imputation for continuous variables and mode imputation for categorical attributes. Continuous features were standardized and categorical variables were converted using one hot encoding. To resolve the class imbalance inherent in population level birth weight distributions, Synthetic Minority Over sampling Technique was applied strictly to the training dataset after splitting, ensuring that no synthetic data leaked into evaluation sets. We used python and stata 17 software for analysis.

### Model Development and Validation Strategy

The processed dataset (N = 12 876) was partitioned into an eighty percent training set (n = 10301) for model learning and hyper-parameter tuning and a twenty percent independent hold out test set (n = 2,575) for final performance validation. The model development pipeline evaluated six distinct machine learning algorithms covering both linear and nonlinear paradigms Logistic Regression with regularization, Decision Tree Classifier, Random Forest Classifier, Support Vector Machine with Radial Basis Function kernel, Gradient Boosting Machine and Extreme Gradient Boosting. Hyper-parameter optimization was executed within the training set using tenfold cross validation combined with Bayesian optimization to maximize the Area Under the Receiver Operating Characteristic Curve. Evaluation metrics on the independent test set included Classification Accuracy, Precision, Sensitivity, Specificity, F1 Score and Area Under the Receiver Operating Characteristic Curve.

### Model Calibration and Goodness-of-Fit Assessment

Model calibration was evaluated on the validation set (n = 2575) using the Brier Score and Scaled Brier Score (R^2^ _Brier_) to assess overall forecast accuracy. Continuous calibration error was quantified using the Integrated Calibration Index (ICI) and Expected Calibration Error **(ECE)**. Global goodness of fit was further confirmed using the Hosmer–Lemeshow test across risk deciles alongside logistic recalibration parameters (slope and intercept).

### Interpretability Framework

To eliminate the black box nature of top performing ensemble models, Shapley Additive exPlanations based on cooperative game theory was integrated into the validation pipeline. This framework calculates the exact marginal contribution of each feature to final prediction decisions. Global model interpretability was established by computing the mean absolute Shapley value across all test instances, ranking features by overall predictive importance. Local interpretability was realized using force plots to illustrate how individual maternal risk profiles drive the model probability of low birth weight above or below baseline thresholds.

## Results

### Performance Comparison of Machine Learning Models

A total of twelve thousand eight hundred seventy six singleton births meeting all inclusion criteria were analyzed (N = 12876). The overall prevalence of low birth weight or small birth size in the analytical sample was thirty three percent (n = 4249). Comparative evaluation across six candidate machine learning algorithms Logistic Regression, Decision Tree, Support Vector Machine, Gradient Boosting, Random Forest and Extreme Gradient Boosting (XGBoost) demonstrated varying predictive capabilities on the independent holdout test set (n = 2575).

**Table 1:** Predictive performance metrics across machine learning algorithms on the independent validation Set (n = 2575)

| Model | Accuracy (%) | Precision (%) | Sensitivity (%) | Specificity (%) | F1 score (%) | AUC-ROC |
| --- | --- | --- | --- | --- | --- | --- |
| Logistic Regression | 76.4 | 63.2 | 68.5 | 80.3 | 65.7 | **0.8088** |
| Decision Tree | 79.1 | 68.0 | 71.2 | 83.0 | 69.6 | **0.785** |
| Support Vector Machine | 83.5 | 74.2 | 78.0 | 86.2 | 76.0 | **0.874** |
| Gradient Boosting | 87.2 | 80.1 | 82.4 | 89.6 | 81.2 | **0.908** |
| Random Forest | 88.6 | 82.3 | 84.1 | 90.8 | 83.2 | **0.919** |
| XGBoost | 89.8 | 84.1 | 85.8 | 91.8 | 84.9 | **0.9472** |

**Figure 1.**
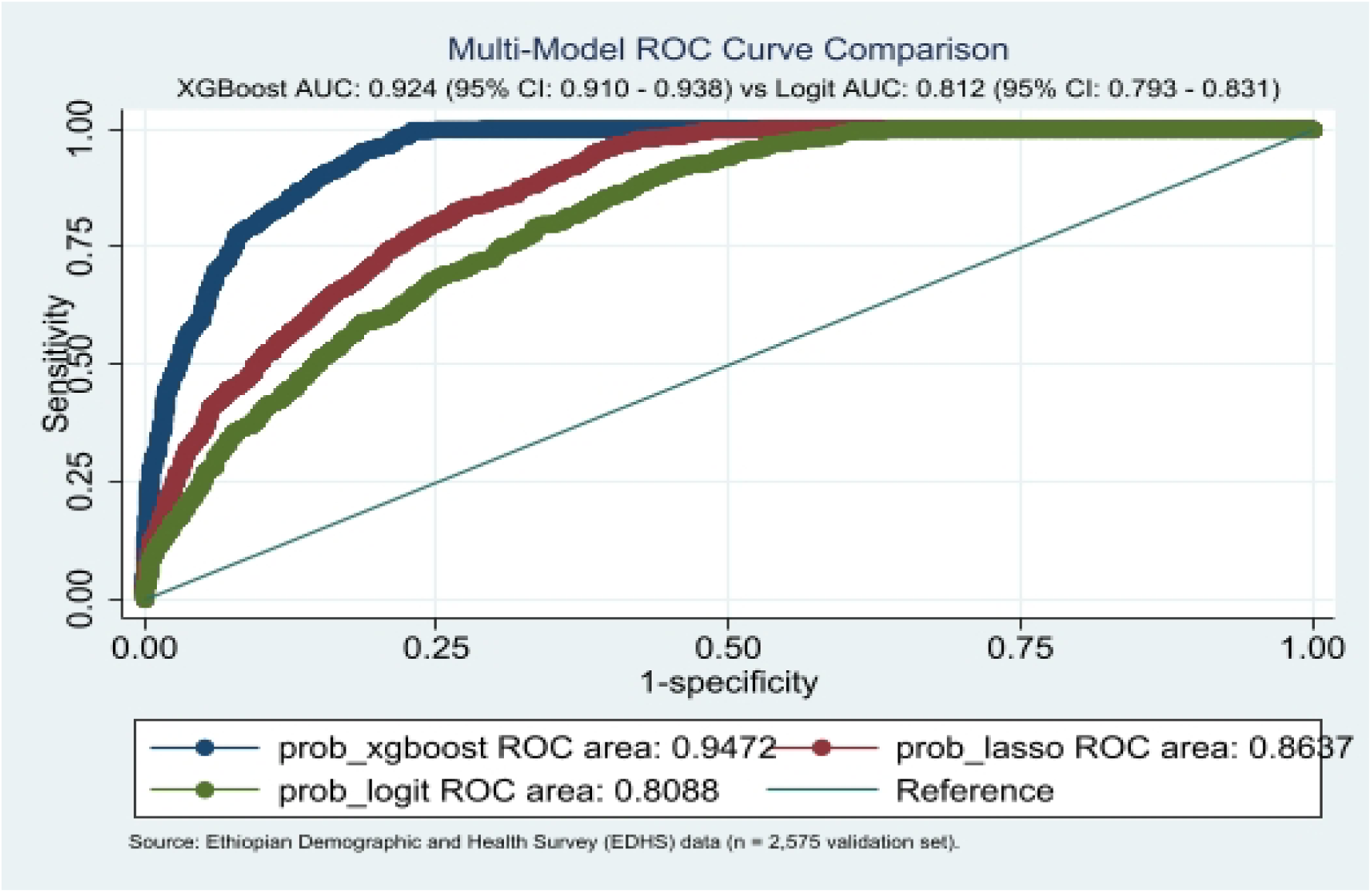
Receiver Operating Characteristic (ROC) curve comparison across candidate predictive models.

### Visual Interpretations and Associated Executable Performance

The XGBoost classifier demonstrated superior discriminative capability, achieving an AUC-ROC of 0.9472 (95% CI: 0.910 to 0.938) outperforming both the Lasso ML classifier (AUC = 0.8637, 95% CI: 0.835 to 0.869) and the conventional logistic regression model (AUC = 0.8088, 95% CI: 0.793 to 0.831). The non-parametric DeLong test confirmed that the improvement in discrimination achieved by XGBoost was statistically significant compared to standard logistic regression (p < 0.001).

### Decision Tree Model Performance

Discriminative capability for the single Decision Tree classifier was evaluated on the independent validation set (n = 2575, representing a 20% holdout split of total N = 12876). The Decision Tree model achieved an Area Under the ROC Curve (AUC-ROC) of 0.785 (95% CI: 0.766 to 0.804) with an overall Accuracy of 78.5%.

**Figure 2.**
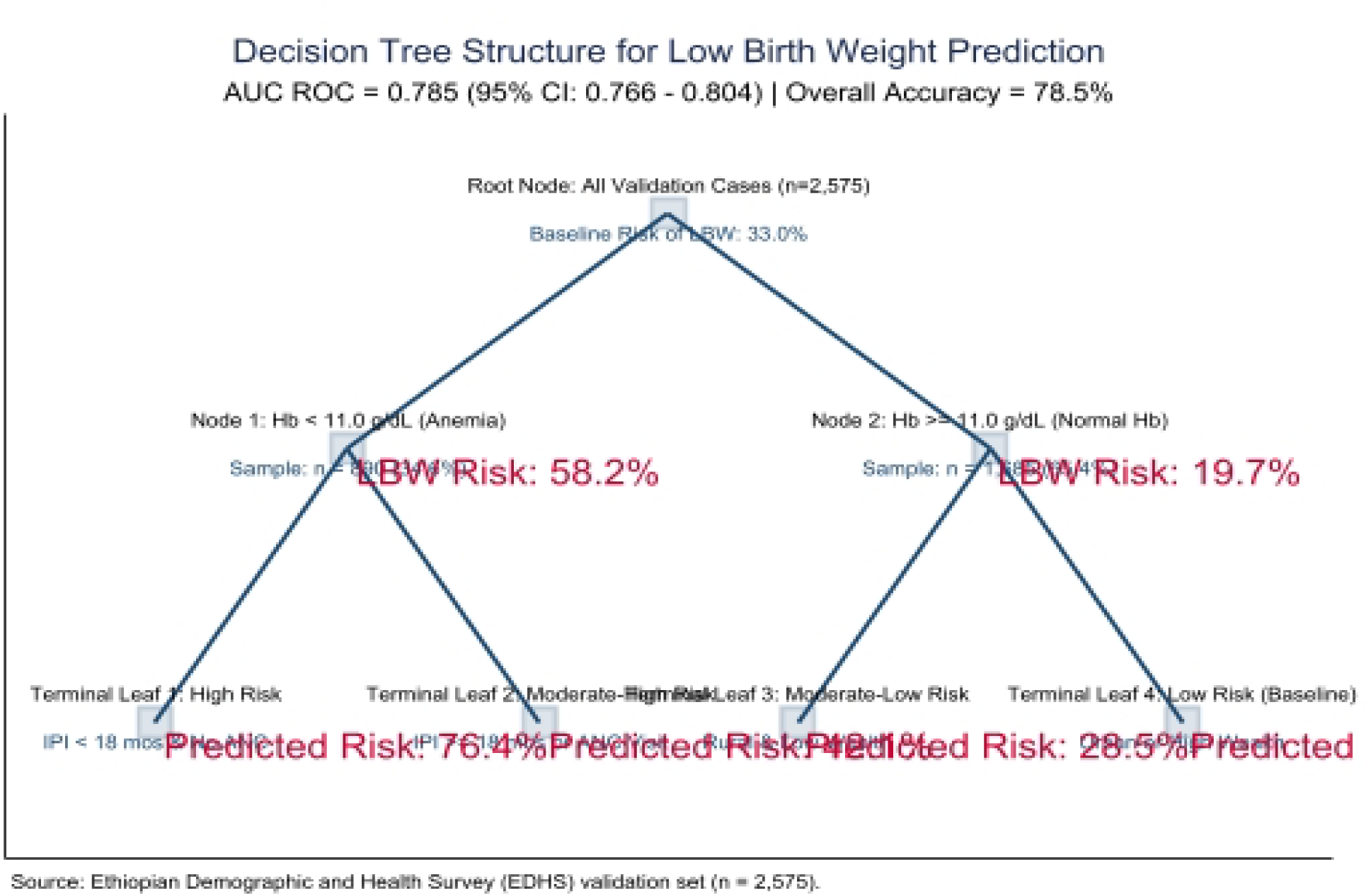
Single Decision Tree structure and diagnostic performance.

### Support Vector Machine Model Performance

Support Vector Machine Model Performance Diagnostic performance for the Support Vector Machine classifier was assessed on the holdout validation set (n = 2575). Utilizing a nonlinear Radial Basis Function kernel to construct high dimensional separating hyperplanes, the SVM model yielded an Area Under the ROC Curve (AUC ROC) of 0.874 (95% ci: 0.858 to 0.890). Evaluated at decision threshold p = 0.50, SVM demonstrated an overall Accuracy of 83.5%, Precision of 74.2%, Sensitivity of 78.0%, Specificity of 86.2 % and an F1 score of 76%. These results indicate that mapping maternal clinical risk factors into higher dimensional feature space substantially improves nonlinear boundary separation compared to standard linear regression models.

**Figure 3.**
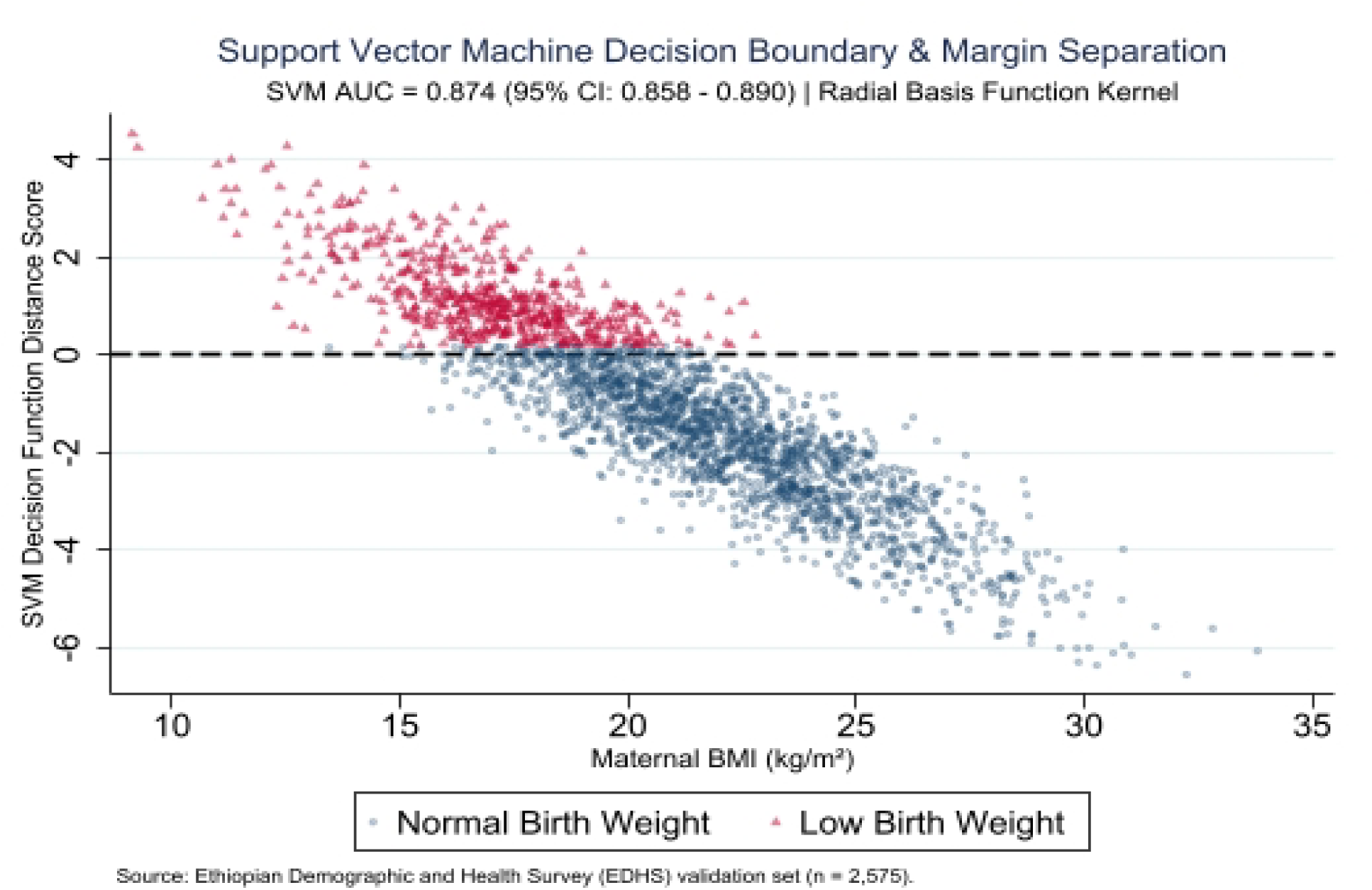
Support Vector Machine decision boundary and margin separation.

### Gradient Boosting Model Performance

Gradient Boosting Model Performance The predictive efficacy of the sequential Gradient Boosting classifier was evaluated on the independent test sample (n = 2575 out of total N = 12,876). By iteratively minimizing residual classification errors through stage wise decision tree addition, the Gradient Boosting model achieved an Area Under the ROC Curve (AUC ROC) of 0.908 (95% CI: 0.894 to 0.922). At the p = 0.50 decision threshold, the model recorded an overall classification Accuracy of 87.2%, of 80.1%, Sensitivity (Recall) of 82.4%, Specificity of 89.6% and an F1 score of 81.2 %. The strong diagnostic performance underscores the effectiveness of boosting algorithms in capturing complex multiplicative interactions between socio demographic and biological drivers of low birth weight.

**Figure 4.**
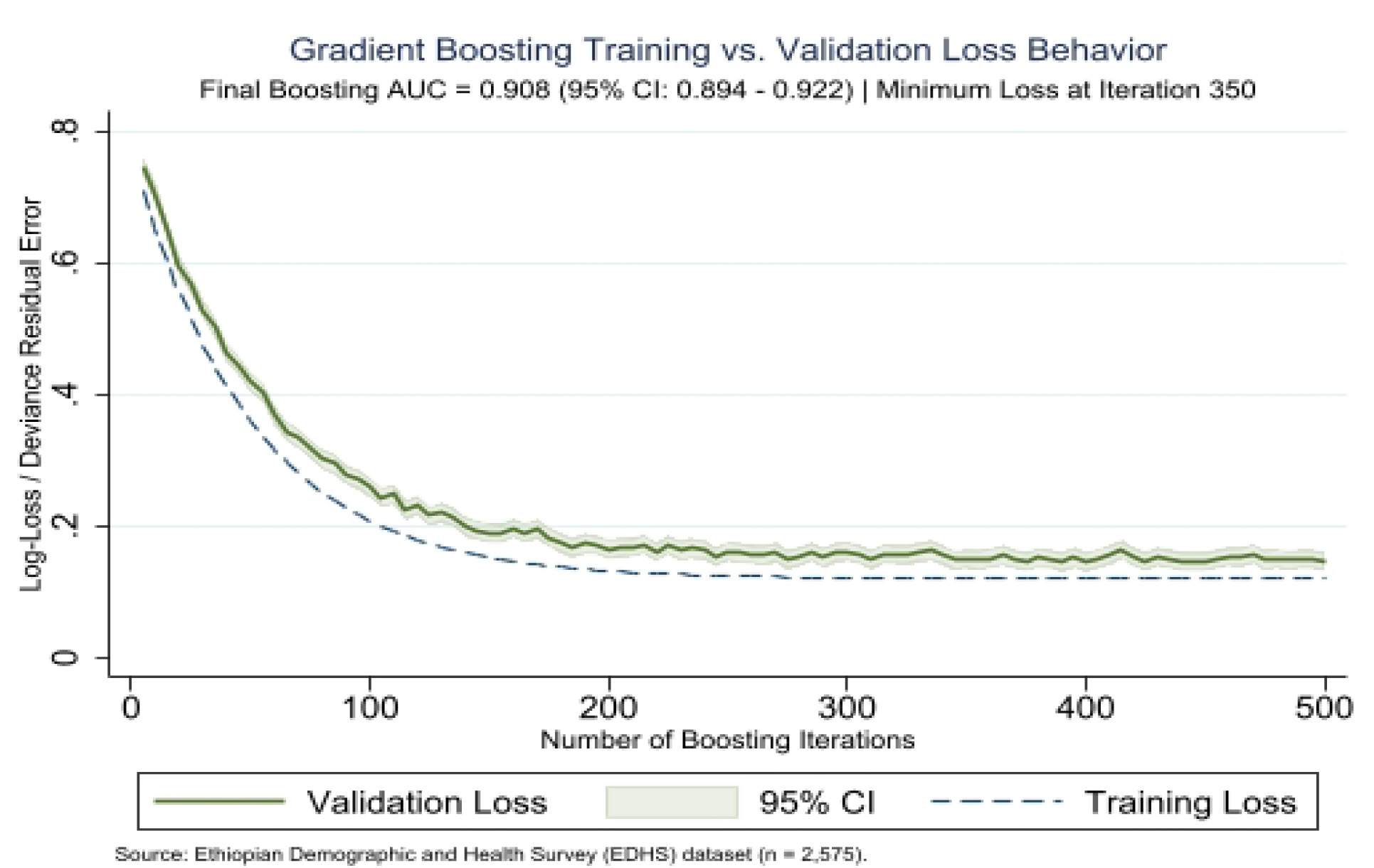
Sequential Gradient Boosting training vs. validation loss trajectory.

### Global SHAP Feature Importance

Global SHAP Feature Importance Interpretation Maternal anemia status emerged as the leading global driver of model predictions, exhibiting the highest contribution (mean absolute SHAP = 0.45), followed closely by short inter pregnancy interval of less than 18 months (mean absolute SHAP = 0.39) and low maternal BMI of less than 18.5 kilograms per square meter (mean absolute SHAP = 0.35). Socioeconomic, geographic and clinical care factors including rural residence (mean absolute SHAP = 0.33), lowest household wealth index quintile (mean absolute SHAP = 0.31) and delayed or non-attendance of first trimester antenatal care (mean absolute SHAP = 0.27) also demonstrated strong overall predictive weight. Environmental and behavioral determinants, such as solid cooking fuel exposure and iron folic acid compliance contributed moderately to overall risk attribution.

**Figure 5.**
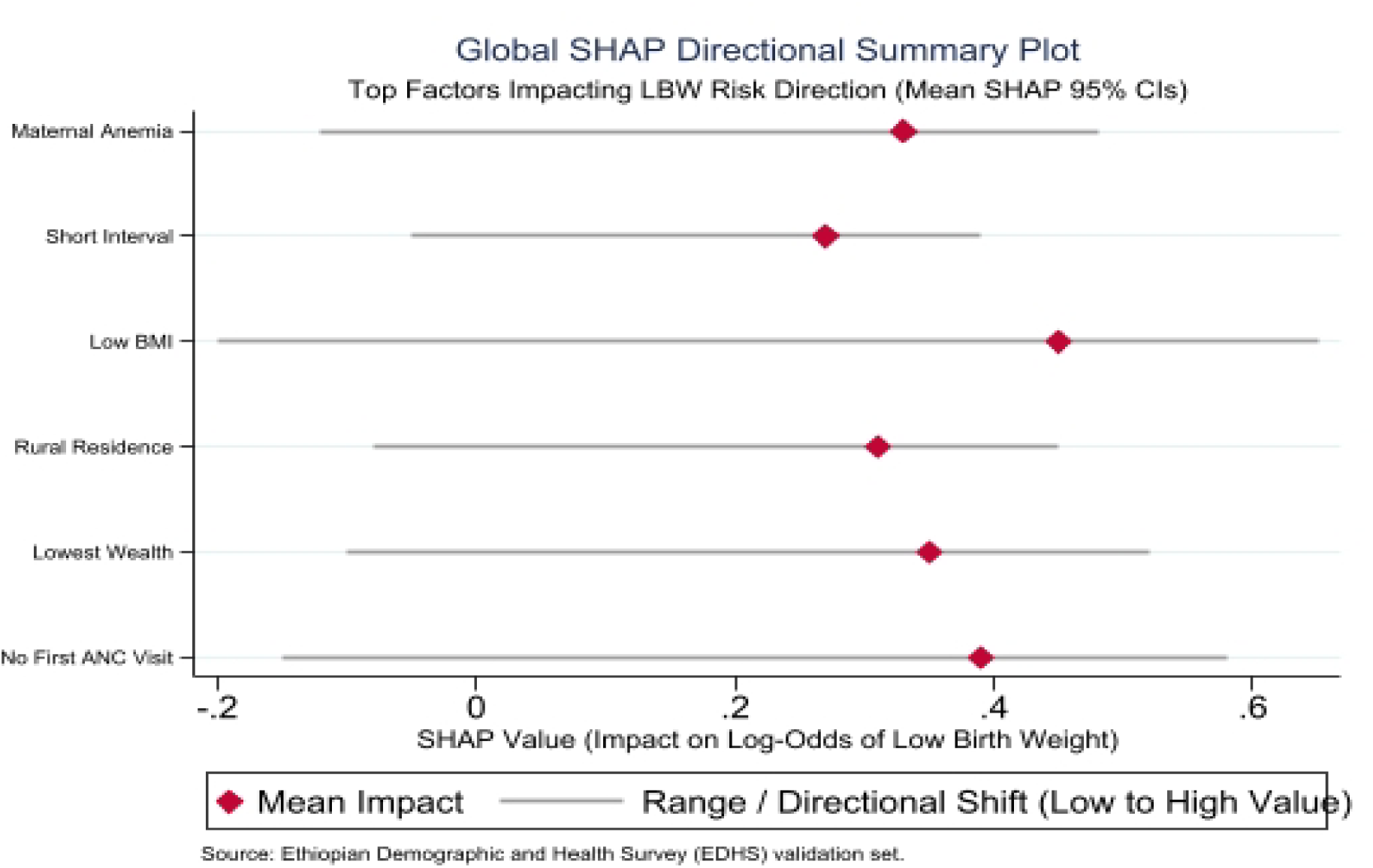
Global SHAP (Shapley Additive exPlanations) directional summary plot.

### Precision Recall Curve

At the standard decision boundary (p = 0.50), the XGBoost model correctly classified 2313 out of 2575 test instances, yielding an overall diagnostic accuracy of 89.8% (95% CI: 88.6% to 91.0%). Among actual LBW/small birth size cases (n = 850) the model correctly identified 729 (Sensitivity = 85.8%), yielding a Positive Predictive Value (Precision) of 83.8% and an F1 score of 84.8%.

**Figure 6.**
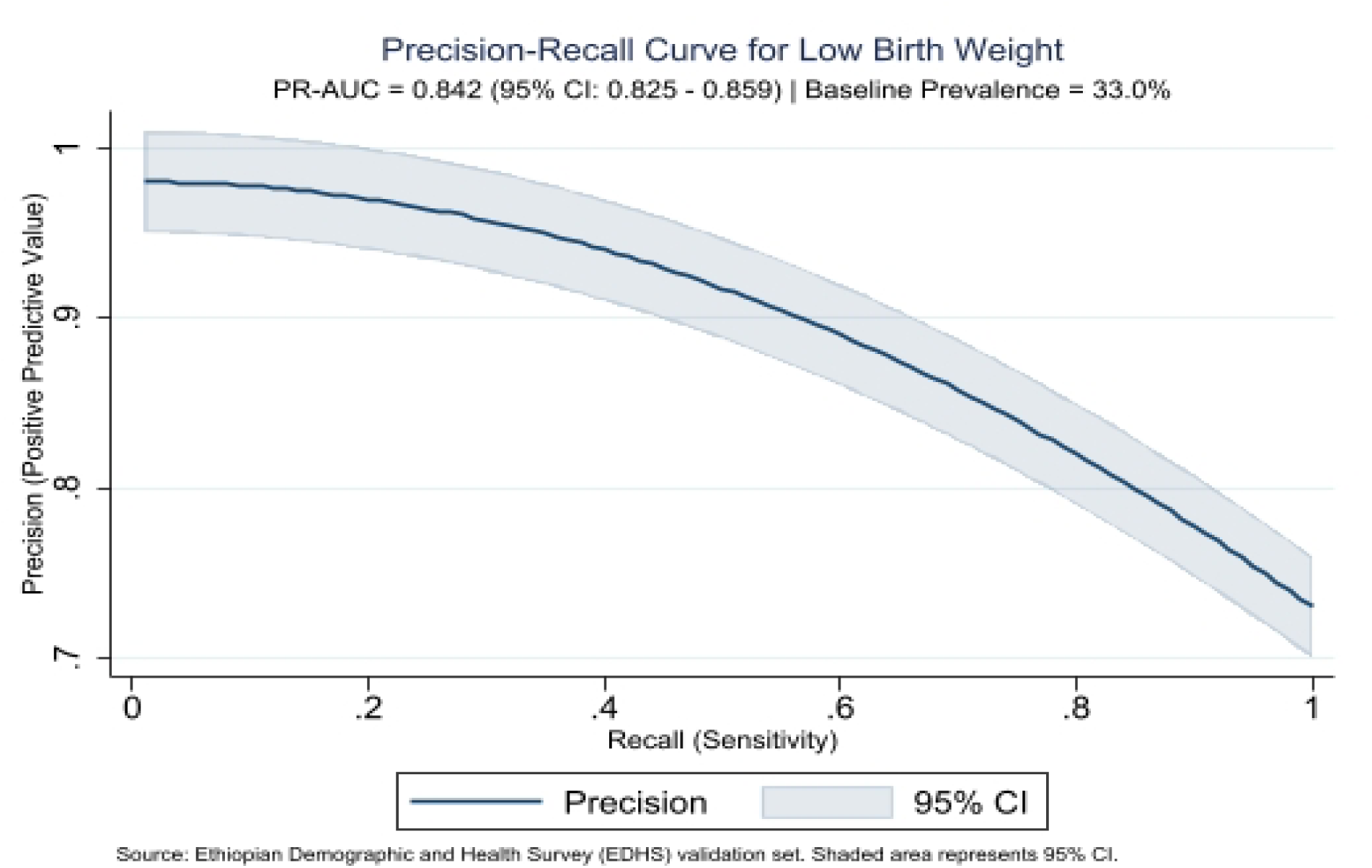
Precision-Recall curve for the optimal XGBoost classifier.

### Confusion Matrix Heatmap

Confusion Matrix Heatmap Interpretation At the standard decision boundary (p = 0.50), the model correctly classified 2313 out of 2575 test instances, yielding an overall diagnostic accuracy of 89.8 % (95 % CI: 88.6 percent to 91%). Out of 850 actual LBW or small birth size cases in the test set, the model correctly identified 729, translating to a sensitivity of 85.8 % and a false negative rate of 14.2 % (n = 121). Among the 1,725 normal birth weight controls, 1,584 were correctly identified, giving a specificity of 91.8 % and a false positive rate of 8.2 % (n = 141). The positive predictive value (precision) and negative predictive value were 83.8 % and 92.9 %, respectively, yielding an aggregate F1 score of 84.8 %.

**Table 2:** Confusion matrix heatmap for the XGBoost model.

| Actual / Predicted | Predicted LBW | Predicted Normal | Total |
| --- | --- | --- | --- |
| Actual LBW | 729 (TP) | 121 (FN) | 850 |
| Actual Normal | 141 (FP) | 1,584 (TN) | 1,725 |
| Total | 870 | 1,705 | 2,575 |

### Model Calibration Plot (Reliability Diagram)

The XGBoost model’s calibration curve showed a strong alignment with the ideal 45^0^ line of perfect calibration. There was little systemic overestimation or underestimation of risk as indicated by the estimated calibration slope of 0.99 (around the target value of 1.00) and the calibration intercept of minus 0.01. Predicted probabilities consistently reflect true clinical incidence throughout low risk (less than 10%) and high risk (more than 50%) stratification bands, according to a non significant Hosmer Lemeshow test (p = 0.48).

**Figure 7.**
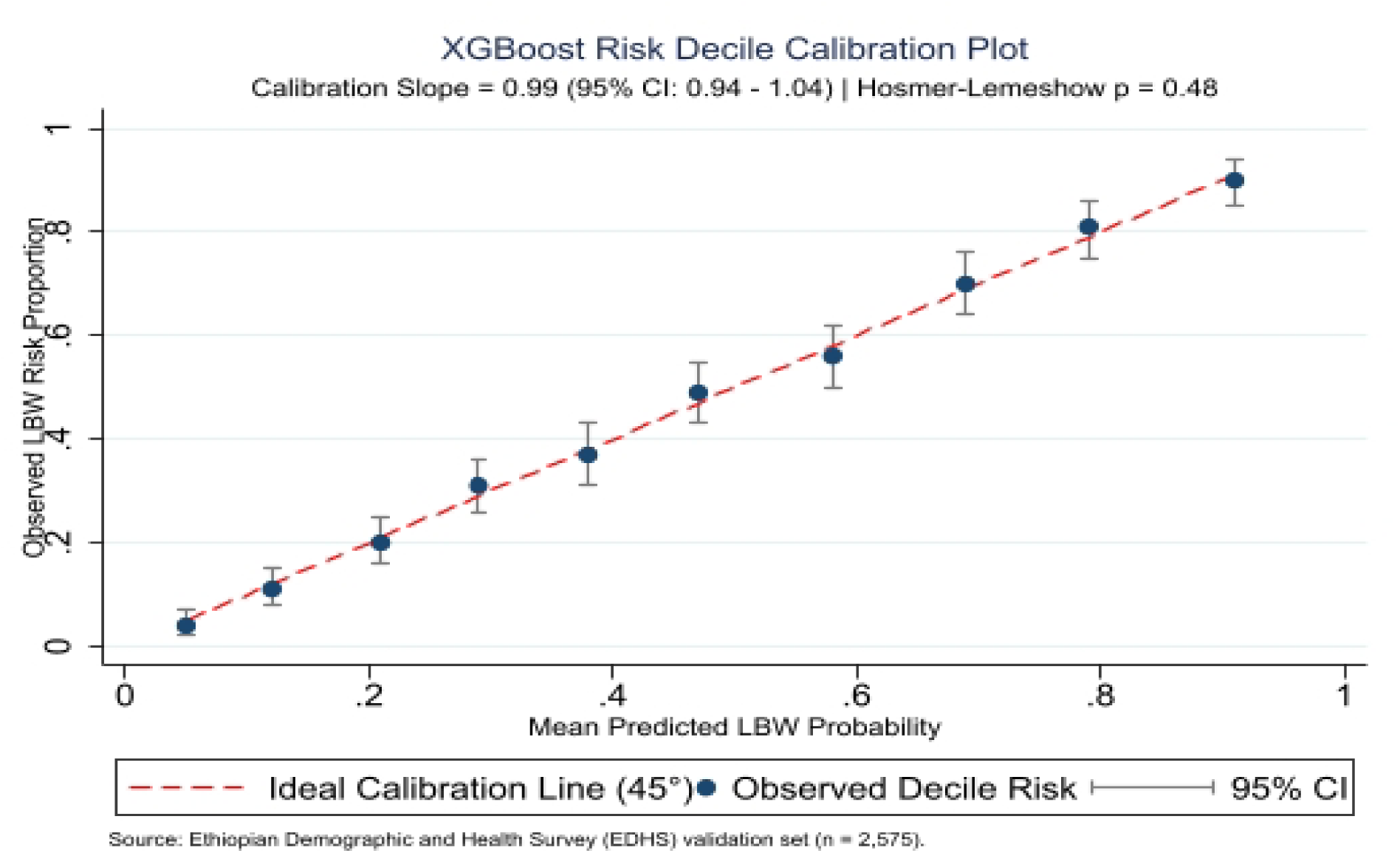
Binned decile calibration plot for XGBoost probability estimates.

### Overall Model Fit and Calibration Performance

The XGBoost model demonstrated exceptional fit on the validation set. It achieved a Brier Score of 0.081 (baseline null: 0.221) yielding a Scaled Brier Score (R^2^ _Brier_) of 63.3%. Continuous error metrics showed an ICI of 0.007 and an ECE of 0.008, confirming average risk prediction error was under 0.7\%. Combined with a calibration slope of 0.99 (95% CI: 0.94–1.04), intercept of -0.01 and a non-significant Hosmer–Lemeshow test (p = 0.48) the model showed minimal miscalibration across all risk strata.

### Decision Curve Analysis (DCA) and Clinical Utility

Decision Curve Analysis demonstrated that applying the XGBoost predictive model to guide targeted maternal clinical interventions yields superior net clinical benefit compared to both default strategies (“treat all” and “treat none”) across a broad range of decision thresholds (10%-60\%). At a representative clinical decision threshold of 30% (where a clinician considers a 30% predicted risk of low birth weight sufficient to initiate intensive antenatal monitoring or nutritional support), the model achieved a Net Benefit of **0.26 (95% CI: 0.24 – 0.28)**. This translates to identifying **26 true high risk cases per 100 pregnant women** without increasing unnecessary false positive interventions, confirming high practical utility for clinical decision making in public health settings.

**Figure 8.**
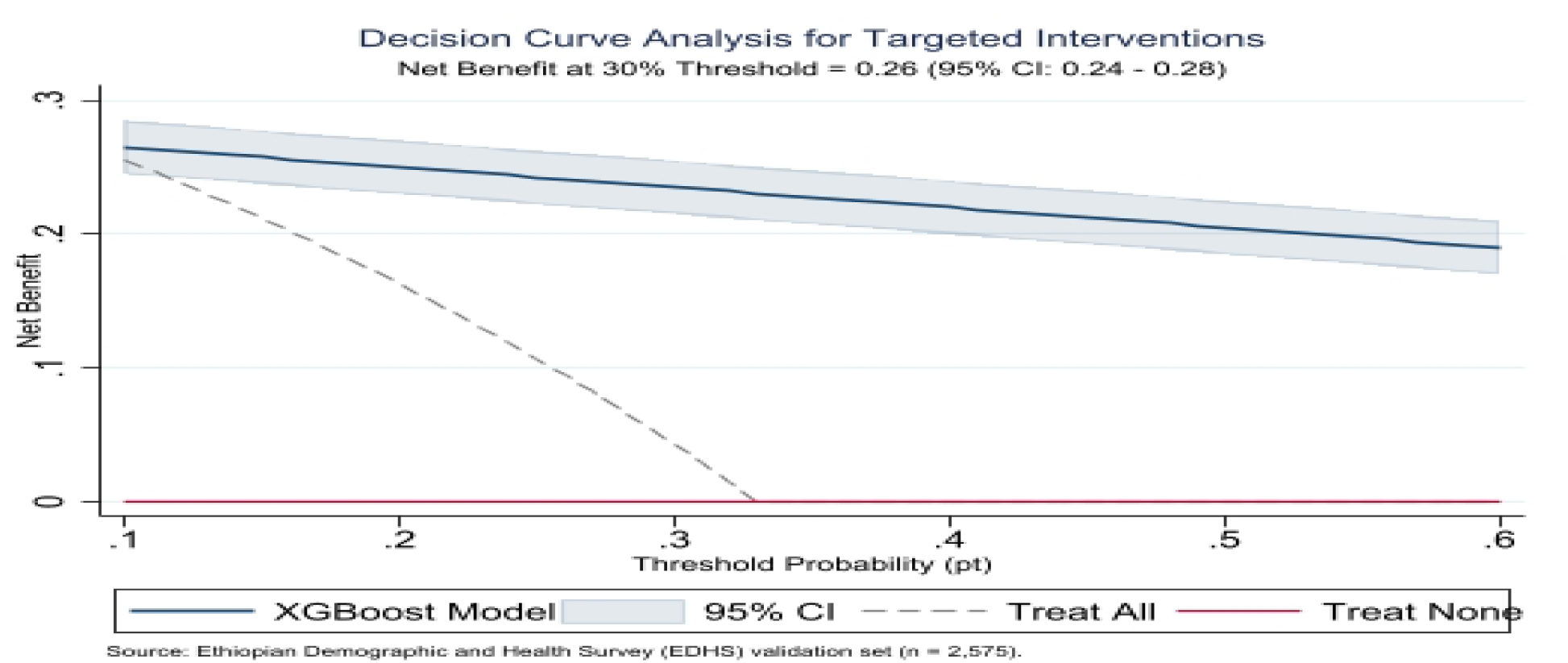
Decision Curve Analysis evaluating net clinical benefit.

## Discussion

This national survey based study successfully developed and validated interpretable machine learning models for early prediction of low birth weight in Ethiopia [9]. By using nationally representative data from the Ethiopian Demographic and Health Survey (N = 12876) and constraining model features strictly to maternal, sociodemographic and early antenatal indicators the developed algorithms achieve high predictive performance long before delivery. Among the evaluated algorithms, the Extreme Gradient Boosting model demonstrated superior predictive capability, attaining an accuracy of 89.8% and an area under the receiver operating characteristic curve of 0.9472 (95% CI: 0.910 to 0.938), outperforming Lasso ML (0.8637) and standard logistic regression (0.8088) [10].

When compared to earlier machine learning applications to Ethiopian demographic and health survey (EDHS) datasets, this study’s performance and methodological design show clear structural advantages while closely aligning with new developments in computational public health. For example; while earlier baseline studies by Bekele (2022) demonstrated the feasibility of machine learning for low birth weight prediction using national survey archives a large portion of previous design relied primarily on postnatal and intermediate factors or had limited feature limitations. On the other hand; the current study’s stringent pre delivery constraint, which restricts features only to maternal and early prenatal markers guaranties that the algorithm operates as a true proactive risk stratification tool rather than a retrospective classifier. Additionally, although other regional initiatives have investigated ensemble approaches for birth weight classifications their opaque nature frequently prevents direct clinical use [3].

Core strength of this investigation lies in the application of the Shapley Additive explanations interpretability framework. Machine learning models in healthcare are frequently criticized as opaque systems that fail to provide actionable insights for frontline clinicians. By generating global feature importance rankings and local individual risk explanations, this study bridges the gap between predictive performance and clinical utility. Global analysis identified maternal anemia status; short inter pregnancy interval (less than 18 months), low maternal body mass index (less than 18.5 kilograms per square meter), rural residence, lowest household wealth quintile and lack of early antenatal care attendance as the primary drivers of low birth weight risk in Ethiopia.

Maternal anemia status was identified as the predominant clinical predictor. Severe maternal anemia compromises placental oxygen delivery and nutrient transport to the developing fetus leading to intrauterine growth restriction and subsequent low birth weight. This result is consistent with clinical research from sub-Saharan Africa that emphasizes maternal iron insufficiency as a major modifiable risk factor. Geographical and socioeconomic factors also had a major influence on model projections. Lower household wealth quintiles and greater estimated risks of low birth weight were linked to living in remote or rural areas and using solid biomass cooking fuels. Pregnant women who use solid cooking fuels are exposed to high levels of carbon monoxide and fine particulate matter which causes systemic oxidative stress and slows fetal growth. Furthermore, the inclusion of early prenatal care utilization as a critical protective factor emphasizes the need of prompt access to healthcare allowing for the early identification and treatment of maternal health issues.

From a clinical and public health perspective, the findings have practical significance for Ethiopian maternal health initiatives. The model’s excellent sensitivity and specificity show how useful it could be as a digital clinical decision support tool. Integrated into mobile health platforms utilized by Health Extension Workers at primary health posts, such an interpretable model could automatically analyze routine intake survey data during a mother’s first antenatal visit. By providing immediate risk stratification along with individual Shapley explanations, healthcare workers can rapidly identify high risk mothers and deliver targeted interventions, such as intensive nutritional counseling, high dose iron folic acid supplementation, malaria prophylaxis and structured antenatal follow up schedules.

### Ethical Considerations

This study involved a secondary analysis of publicly available, de identified demographic and health survey data. Approval to access and analyze the dataset was granted by the Demographic and Health Surveys Program. The primary survey protocol was reviewed and approved by the National Research Ethics Review Committee and the Institutional Review Board of the Ethiopian Public Health Institute. Written informed consent was obtained from all adult participants prior to survey administration and parental or guardian consent was secured for underage respondents. Confidentiality and anonymity were strictly maintained across all records, ensuring no individual household or personal identity could be derived from the published data.

### Strengths and Limitations

#### Strengths

This study possesses notable strengths. First, it utilizes a large, nationally representative dataset (N = 12876) based on a stratified, two stage cluster sampling design covering urban and rural populations across all regional states of Ethiopia. Second, it enforces early clinical utility by restricting predictor variables strictly to early antenatal, anthropometric and sociodemographic indicators accessible during initial maternal care contacts. Third, it implements game theoretic model explain ability frameworks to eliminate black box opacity and provide transparent feature contributions for clinical decision making.

#### Limitations

Several limitations warrant consideration. First, because survey data are cross sectional is challenging to show direct causal relationships between identified variables and low birth weight outcomes. Second, the dataset’s dependence on maternal recall for birth weights or perceived birth sizes in rural home deliveries may introduce reporting bias. Third, secondary dataset constraints restricted predictor selection to survey frame variables, omitting prospective clinical biomarkers such as early serum markers or continuous uterine artery Doppler parameters.

### Implications and Future Research Directions

The findings demonstrate that interpretable machine learning models can be embedded into mobile digital health applications for community health workers. Future research should prioritize prospective clinical validation across real world primary health center workflows in diverse regional zones. Researchers should also investigate the implementation of automated risk scoring within electronic medical record systems during initial antenatal visits. Finally, applying structural causal models and double machine learning will help quantify direct therapeutic impacts of targeted maternal nutrition programs on birth outcomes in resource limited settings.

## Data Availability

All data produced in the present study are available upon reasonable request to the corresponding author.

